# Greater Role of Cognitive Impairment Over Fatigue in Post-COVID-19 Quality of Life: A Post-Hoc Analysis of a Randomized Controlled Trial

**DOI:** 10.1101/2024.03.20.24304411

**Authors:** Angela T.H. Kwan, Moiz Lakhani, Gia Han Le, Gurkaran Singh, Kayla M. Teopiz, Ziji Guo, Felicia Ceban, Kanwarpreet Kaur Dhaliwal, Sebastian Badulescu, Roger Ho, Taeho Greg Rhee, Bing Cao, Giacomo d’Andrea, Roger S. McIntyre

## Abstract

**Background:** Post COVID-19 Condition (PCC) is a common and debilitating condition with significant reports of fatigue and psychosocial impairment globally. The extent to which cognitive symptoms and fatigue contribute to reduced quality of life in affected individuals remains clear.

**Methods:** This is a post-hoc analysis of a randomized, double-blind, placebo-controlled clinical trial that evaluated the effect of vortioxetine on cognitive function in adults with PCC. The post-hoc analysis herein aimed to determine the overall effect of baseline cognitive function [as measured by the Digit Symbol Substitution Test (DSST)] and baseline fatigue severity [as measured by the Fatigue Severity Scale (FSS)] on baseline health-related quality of life (HRQoL) [as measured by the 5-item World Health Organisation Well-Being Index (WHO-5)].

**Results:** A total of 200 participants were enrolled in the primary trial. Due to missing baseline data, our statistical analysis included baseline measures of 147 individuals. Our generalized linear model analysis revealed a significant positive correlation between DSST-measured objective cognitive function and self-reported WHO-5-measured HRQoL (β = 0.069, 95% CI [0.006, 0.131], *p* = 0.032). In contrast, our analysis revealed a significant negative correlation between FSS and WHO-5 scores (β = -0.016, 95% CI [-0.021, –0.011], *p* < 0.001). The beta-coefficient ratio (β_DSST_ / β_FSS_ = 0.069 / 0.016) is calculated as 4.313.

**Conclusions:** Overall, we observed that increased cognitive function was associated with increased HRQoL at baseline in adults with PCC. Moreover, we observed that increased severity of fatigue symptoms was associated with decreased HRQoL at baseline in adults with PCC. Furthermore, we observed that an improvement in cognitive function would have a four-fold greater impact on HRQoL than the effect generated by improvement in fatigue.

## INTRODUCTION

Post COVID-19 condition (PCC) is a widespread, debilitating, and severe phenomenon.^1^ The World Health Organization (WHO) defines PCC as a condition that occurs three months following confirmed coronavirus disease 2019 (COVID-19) infection, even after the acute infection resolves, and cannot be attributed to another diagnosis.^2^ Approximately 10-20% of persons infected with COVID-19 meet the criteria for PCC.^3,4^ PCC is a heterogeneous group of disorders involving multiple organ systems.^5^ Additionally, cognitive impairment (e.g., brain fog) and fatigue are two of the most common features of PCC.^1,6^ Approximately 32% of individuals who experienced PCC reported fatigue, and 22% reported cognitive decline.^1^

Cognitive impairment and fatigue are relevant to the health-related quality of life (HRQOL) and functioning of individuals with PCC; however, the extent of contribution from each domain is under-studied.^1,7,8^ Understanding their respective contributions to HRQOL holds implications for early interventions, the discovery and development of treatments, clinical care planning, and the characterization of mechanistic models in PCC. In this post-hoc study of a randomized controlled trial, we sought to determine the respective impacts of objective cognitive function (e.g., motor speed, attention, and visuoperceptual functions) and fatigue on measures of HRQOL in individuals with PCC.

## MATERIALS AND METHODS

### Study Design and Participants

Herein we conducted a post hoc analysis of a randomized, double blind, placebo-controlled clinical trial investigating the efficacy of vortioxetine in the treatment of cognitive symptoms in adults with PCC. The data and methodology reported herein was obtained from the primary trial which is published elsewhere.^9^ The primary trial was conducted in accordance with the principles of Good Clinical Practice and the Declaration of Helsinki.^10,11^ The primary trial was registered on Clinicaltrials.gov (NCT05047952). The trial design was approved by Advarra, a local research ethics board (REB), that complies with Health Canada regulations, (IRB #00000971). Study recruitment took place in Canada from November 2021 to January 2023. Methods of participant recruitment included media advertisements (e.g., Facebook, Twitter, Instagram, and/or print) or referrals by medical practitioners. Written informed consent was obtained before participation from all eligible participants.

The eligibility criteria for the primary trial included: persons aged ≥ 18 years who were residing in Canada at the time of the study, able to provide documentation of WHO-defined PCC symptoms (i.e., symptoms of COVID-19 occurring within 3 months of acute SAR-CoV-2 infection and persisting for at least 2 months after resolution of acute infection with no other explanation), able to provide documentation of a positive severe acute respiratory syndrome coronavirus 2 (SARS□CoV□2) test (i.e., antigen, serology, and/or PCR). Alternatively, participants without a documented positive SARS-CoV-2 test who were determined to have a probable SARS-CoV-2 infection (such as a prior diagnosis of acute COVID-19 from a healthcare provider or a clinical diagnosis from the study physician) were also deemed eligible. Moreover, participants were required to provide written informed consent at the time of screening/baseline in order to be eligible for the study.

Eligible participants that were using other antidepressants at the time of the study were instructed by the study physician to cease their antidepressant medication for a minimum of 2-4 weeks before the baseline assessment. Additionally, eligible participants were informed via consent forms that the safety and/or efficacy effects of combining the study medication with another antidepressant remains investigational. Individuals who met any of the exclusionary criteria were excluded from study participation **(Supplementary Materials, Table S1)**.

### Procedures

For the duration of an 8-week double blind treatment, 149 eligible participants were randomized (1:1) to receive vortioxetine (5-20 mg/d) or placebo. From weeks 1-2, participants in the vortioxetine group aged 18-65 received vortioxetine at 10 mg/d and participants aged 65+ in this group received vortioxetine at 5 mg/d. From weeks 3 to 8, participants aged 18-65 in the vortioxetine group received vortioxetine at 20 mg/d, and participants in this group aged 65+ received 10 mg/d. The vortioxetine and placebo capsules were identical in size and appearance.

Study visits occurred at baseline and weeks 2, 4, and 8. Due to public health recommendations in Canada during the time of the study, study visits were made available remotely via telephone or online platform (i.e., Zoom) to increase participant retention. Study visits were also made available in-person if preferred or required. A secure online platform (i.e., Ontario Telemedicine Network) was used to conduct visits with the study physician. A safety visit during weeks 8 and 10 was offered to all participants. Participants who withdrew prior to study completion were scheduled for a follow up appointment at the earliest available date.

### Outcome Measures

The primary outcome measure in the primary clinical trial was the effect of vortioxetine on cognitive performance as measured by the Digital Symbol Substitution Test (DSST) Pen/Paper Version and Online CogState Version as part of the CogState Online Cognitive Battery.^9^ The pen-paper version of the DSST was not completed by remote participants. At baseline, and weeks 2 and 8, the DSST was administered.

Additionally, multiple secondary outcome measures were collected in the primary trial, including fatigue as measured by the Fatigue Severity Scale (FSS) and health-related quality of life as measured by the 5-item World Health Organisation Well-Being Index (WHO-5). The FSS and WHO-5 were measured at baseline and weeks 2, 4 and 8. A complete list of all secondary outcome measures of the primary trial are reported in the primary paper.^9^

### Statistical Analysis

All statistical computations were performed utilizing IBM SPSS Statistic software version 28.0.1.1 (15), with a significance level set at α = 0.05. Descriptive statistics were expressed as frequency (%) for categorical variables; the mean [standard deviation (SD)] and median were utilized for continuously distributed variables with a normal distribution. Of the 149 enrolled participants, 11 (7.4%) completed the Pen/Paper Version of the DSST only; 78 (52.3%) completed only the CogState Version, and 60 (40.3%) completed both the Pen/Paper and Online CogState Version. For participants that completed both DSST versions, performances on the Pen/Paper and Online CogState Version were highly and significantly correlated (r = 0.588, *p* < 0.001). Further computations were performed using the combined DSST score as not all participants completed both the Pen/Paper and Online CogState Versions. Specifically, the combined DSST scores were based on participants’ Online CogState DSST scores if they completed both Online CogState and Pen/Paper DSST. If the Online CogState DSST was not completed, participants’ Pen/Paper DSST scores were included in the combined DSST scores. An intent-to-treat (ITT) analysis (i.e., including all randomized participants) was employed to assess DSST and FSS total scores at baseline.

To examine the correlation between objective cognition (DSST) and fatigue (FSS) on HRQoL, a generalized linear model (GLM) with a Poisson probability distribution was performed (WHO-5). Potential covariates identified were participant’s age, sex, race, education, type of cognition test, suspected vs. confirmed COVID-19 diagnosis, and major depressive disorder (MDD) diagnosis.

## RESULTS

### Patient Characteristics

Baseline sociodemographic and clinical characteristics are described in **Table 1**. Of the 200 participants who provided signed informed consent, baseline analysis was conducted on data from only 147 participants due to missing information. No significant differences were observed between the groups for baseline characteristics **(Table 1)**.

**Table 1.**
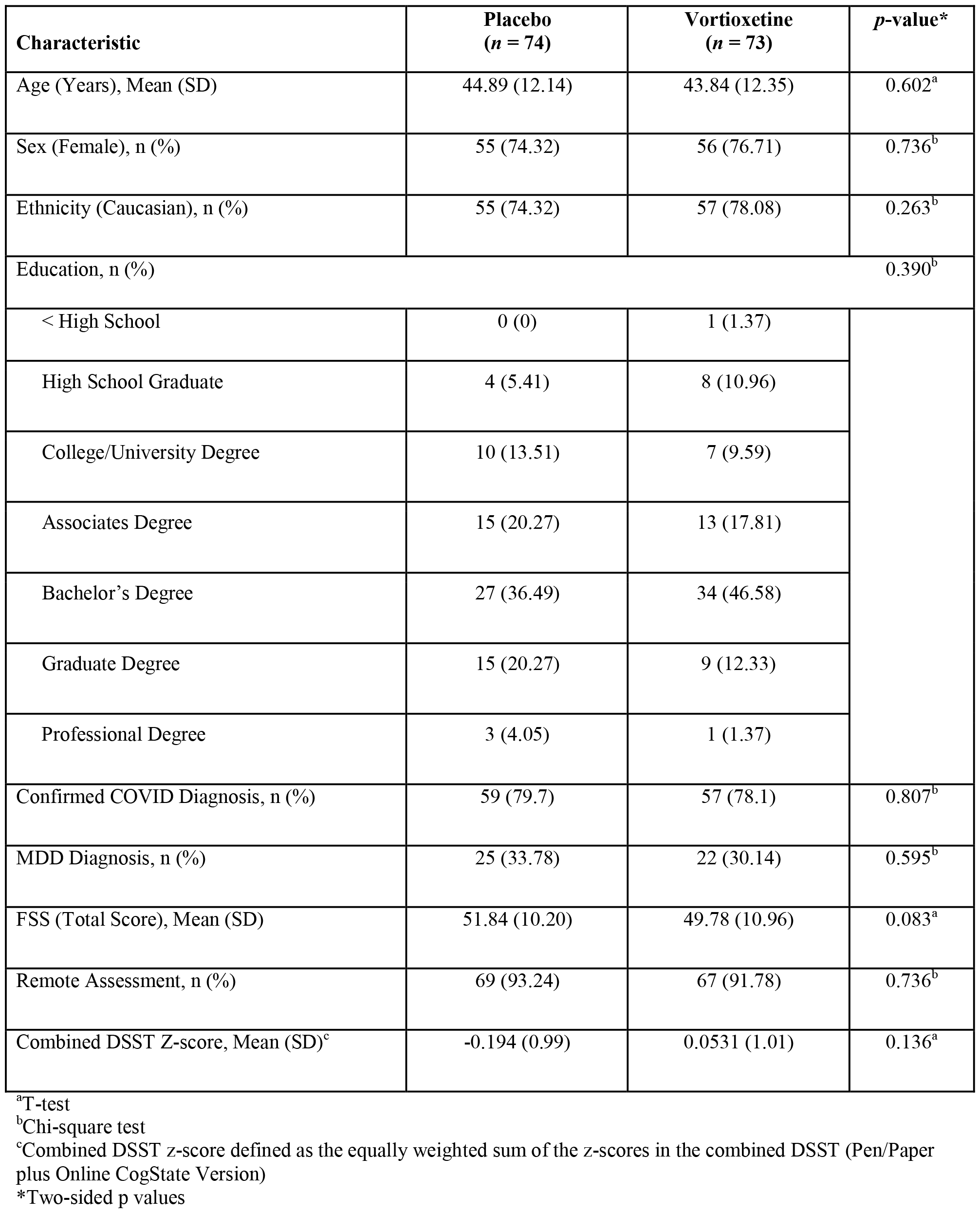
Baseline characteristics of the intent-to-treat population (N = 147).

### Greater Role of Cognitive Function Over Fatigue on Health-Related Quality of Life in Persons with Post-COVID-19 Condition

Results from the GLM analysis demonstrate a significant positive correlation between DSST-measured objective cognitive function and self-reported WHO-5-measured HRQoL (β = 0.069, 95% CI [0.006, 0.131], *p* = 0.032) **(Table 2)**. In contrast, FSS (β = -0.016, 95% CI [-0.021, – 0.011], *p* < 0.001) shows a significant negative correlation with WHO-5 scores **(Table 2)**. The beta-coefficient ratio (β_DSST_ / β_FSS_ = 0.069 / 0.016) is calculated as 4.313. This signifies that an improvement in cognitive function would have a four-fold greater impact on HRQoL than the effect generated by fatigue.

**Table 2.** Generalized linear model of baseline cognition and fatigue on WHO-5 scores.

| IV | Model | $\beta$ | 95% Confidence Interval | | *P*-value |
| --- | --- | --- | --- | --- | --- |
|  |  |  | Lower | Upper |  |
| Combined DSST<sup>a</sup> |  |  |  |  |  |
|  | Combined DSST* | 0.069 | 0.006 | 0.131 | 0.032 |
|  | Age* | -0.001 | -0.006 | 0.004 | 0.660 |
|  | Sex | -0.028 | -0.150 | 0.093 | 0.646 |
|  | Race | -0.019 | -0.043 | 0.004 | 0.111 |
|  | Education* | 0.066 | 0.023 | 0.109 | 0.003 |
|  | Type of Cognitive Test | 0.340 | 0.114 | 0.566 | 0.003 |
|  | Suspected vs. Confirmed COVID-19* | -0.180 | -0.305 | -0.054 | 0.005 |
|  | MDD Diagnosis | 0.074 | -0.038 | 0.185 | 0.196 |
| FSS |  |  |  |  |  |
|  | Intercept | 3.162 | 2.680 | 3.644 | < 0.001 |
|  | FSS | -0.016 | -0.021 | -0.011 | < 0.001 |
|  | Age* | -0.002 | -0.006 | 0.002 | 0.384 |
|  | Sex | -0.076 | -0.199 | 0.047 | 0.225 |
|  | Race | -0.002 | -0.026 | 0.021 | 0.855 |
|  | Education* | 0.056 | 0.013 | 0.099 | 0.011 |
|  | Suspected vs. Confirmed COVID-19* | -0.168 | -0.293 | -0.044 | 0.008 |
|  | MDD Diagnosis | 0.018 | -0.097 | 0.132 | 0.762 |

\* $p < 0.05$ <sup>a</sup>Combined DSST z-score defined as the equally weighted sum of the z-scores in the combined DSST (Pen/Paper plus Online CogState Version)**Abbreviations:** DSST = Digit Symbol Substitution Test; FSS = Fatigue Severity Scale; IV = Independent Variable.

## DISCUSSION

Herein, we found that objective cognitive impairment had a notably greater positive impact on HRQOL in individuals with PCC than fatigue. Both factors significantly contributed to reduced HRQoL, with cognitive impairment exerting a disproportionately larger influence.

It may be conjectured that the greater impact of cognitive impairment on HRQOL stems from the significant involvement of cognition in workplace/academic/social interactions, which is known to be more pronounced in individuals living with PCC.^12,13^ Moreover, such a decline in cognitive function may also lead to attenuated hedonic function, which would be predicted to further decrease HRQoL in persons with PCC. For example, in other disease states such as MDD, it is established that cognitive function—specifically processing speed and executive function—and anhedonic function are dissociable phenomena but also highly overlapping.^14^ Our findings suggest that individuals with PCC should be screened for objective cognitive impairment and fatigue, allowing for a deeper comprehension of their contribution to HRQOL. It is important that treatment efforts correspond to their relative impact on HRQoL, involving strategies to preserve, regain, and protect cognitive function in persons with PCC. Our primary focus should be to address cognitive issues in individuals presenting with this concern.

Several methodological limitations impact interpretations and inferences drawn from our data. First, this was a post-hoc analysis based on data collected during the primary study, and our exploration of the relative contribution of cognitive functioning and fatigue to HRQoL was not pre-determined in the protocol. While we ruled out other medical conditions as the primary cause of presentation, it is possible that prior medical conditions affecting cognitive function might not have been disclosed. Our assessment of HRQoL in PCC also relied on self-reported data from the WHO-5. Different HRQoL assessments and functional neuroimaging measurements of cerebral oxyhemoglobin levels during DSST may yield different results.^15^ Furthermore, our sample was heterogenous and encompassed diverse factors such as acute COVID-19 severity, duration of PCC, number of prior COVID-19 infections, and the number and type of prior vaccinations.

## CONCLUSION

Collectively, our findings show that cognitive impairment has a more pronounced impact on diminishing HRQoL than fatigue. Healthcare practitioners providing care to individuals with PCC should give precedence to therapeutic targets that wield the greatest influence on HRQoL. Furthermore, our data has implications in disease modeling and significantly informs the advancement of therapeutic strategies and development for individuals affected by PCC.

## DATA AVAILABILITY

The data and research materials that support the findings of this study are available from the corresponding author, R.S.M, upon reasonable request and will be anonymized.

