## Supplemental Table 1 for "Greater Role of Cognitive Impairment Over Fatigue in Post-COVID-19 Quality of Life: A Post-Hoc Analysis of a Randomized Controlled Trial"

**SUPPLEMENTARY**

**Table S1.** Exclusion criteria

- Current symptoms were better explained by symptoms of major depressive disorder or bipolar disorder;
- Symptoms were fully explained by pre-existing conditions that may cause cognitive impairment or symptoms similar to those seen in PCC [e.g., attention deficit/hyperactivity disorder (ADHD), major neurocognitive disorder, schizophrenia, chronic fatigue syndrome (CFS)/encephalitis meningitis (EM), as assessed by Mini International Neuropsychiatric Interview (M.I.N.I.) 7.0.2];
- Known intolerance to vortioxetine and/or prior trial of vortioxetine with demonstrated inefficacy;
- Current alcohol and/or substance use disorder, as confirmed by the M.I.N.I. 7.0.2;
- Presence of comorbid psychiatric disorder that is a primary focus of clinical concern, as confirmed by the M.I.N.I. 7.0.2;
- Previous history of mania/hypomania;
- Taking medications approved and/or employed off-label for cognitive dysfunction (e.g., psychostimulants);
- Any medication for a general medical disorder that may affect cognitive function (as per clinical judgment);
- Use of benzodiazepines within 12 hours of cognitive assessments;
- Consumption of alcohol within eight hours of cognitive assessments;
- Any physical, cognitive, or language impairments sufficient to adversely affect data derived from cognitive assessments;
- Diagnosed reading disability or dyslexia;
- Clinically significant learning disorder by history;
- Treatment with electroconvulsive therapy (ECT) in the last 6 months;
- History of moderate or severe head trauma (e.g., loss of consciousness for > 1 hour), other neurological disorders, or unstable systemic medical diseases that are likely to affect the central nervous system (as per clinical judgment);
- Pregnant and/or breastfeeding; received investigational agents as part of a separate study within 30 days of the screening visit;
- Actively suicidal/presence of suicidal ideation or evaluated as being at suicide risk (as per clinical judgment);
- Currently receiving treatment with monoamine oxidase inhibitor (MAOI) antidepressants, antibiotics such as linezolid or intravenous methylene blue;
- Previous hypersensitivity reaction to vortioxetine or any components of the formulation;
- Previously reported angioedema in persons treated with vortioxetine;
- Serotonin syndrome;
- Abnormal bleeding;
- Angle closure glaucoma;
- Hyponatremia;
- Moderate hepatic impairment;
- Active seizure disorder/epilepsy that is not controlled by medication (as per clinical judgment);
- Presence of any unstable medical conditions;
- Inability to follow study procedures;
- And inability to give informed consent.
